# A magnetic bead immunoassay to detect human IgG reactive to SARS-CoV-2 Spike S1 RBD produced in *Escherichia coli*

**DOI:** 10.1101/2021.12.22.21268286

**Authors:** Marcelo S. Conzentino, Ana C. A. Gonçalves, Nigela M. Paula, Fabiane G. M. Rego, Dalila L. Zanette, Mateus N. Aoki, Jeanine M. Nardin, Luciano F. Huergo

**Affiliations:** Setor Litoral, UFPR Matinhos, PR, Brazil; Post-Graduation Program in Pharmaceutical Sciences, UFPR, Curitiba, PR, Brazil; Instituto Carlos Chagas – FioCruz, PR, Brazil; Hospital Erasto Gaertner, Curitiba, PR, Brazil

**Keywords:** COVID-19, magnetic immunoassay, SARS-CoV-2, high throughput, magnetic beads

## Abstract

Immunological assays to detect SARS-CoV-2 Spike receptor binding domain antigen seroconversion in humans are important tools to monitor the levels of protecting antibodies in the population in response to infection and/or immunization. Here we describe a simple, low cost and high throughput Ni^2+^ magnetic bead immunoassay to detect human IgG reactive to Spike S1 RBD Receptor Binding Domain produced in *Escherichia coli*. A 6xHis tagged Spike S1 RBD was expressed in *E. coli* and purified by affinity chromatography. The protein was mobilized on the surface of Ni^2+^ magnetic beads and used to investigate the presence of reactive IgG in the serum obtained from pre-pandemic and COVID-19 confirmed cases. The method was validated with a cohort of 290 samples and an area under the receiver operating characteristics curve of 0.94 was obtained. The method operated with>82% sensitivity at 98% specificity and was also able to track human IgG raised in response to vaccination with Comirnaty with 85% sensitivity. The IgG signal obtained with the described method was well correlated with the signal obtained when pre fusion Spike produced in HEK cell lines were used as antigen. This novel low-cost and high throughput immunoassay may act as an important tool to investigate protecting IgG antibodies against SARS-CoV-2 in the human population.

## Introduction

In December 2019 a novel beta-coronavirus named SARS-CoV-2 was identified and was responsible for the COVID-19 pandemic [1]. At the end of November 2021 260,000,000 cases of COVID-19 have been reported and the disease has caused more than 5,150,000 deaths worldwide https://coronavirus.jhu.edu/map.html (accessed 21/11/2021). The cycle of SARS-CoV-2 infection stars with the interaction between the Receptor-Binding-Domain (RBD) of the SARS-CoV-2 Spike protein and the human receptor ACE2 [2]. Several studies indicate that human antibodies reactive to SARS-CoV-2 Spike, especially those elicited against the Spike RBD, can neutralize virions particles by blocking the interaction between Spike with the human receptor ACE2 [3], [4]. Hence, a method enabling the identification and quantification of human antibodies reactive to SARS-CoV-2 Spike and Spike RBD can act as an important toll to monitor the level of protecting antibodies to SARS-CoV-2 within the human population.

Since the beginning of the COVID-19 pandemic, several immunological assays were developed and employed to detect human antibodies against SARS-CoV-2 Spike [5], [6], [7], [8]. These immunoassays typically use recombinantly expressed and highly purified Spike or Spike RBD to investigate antibodies in human samples. The production of recombinant Spike antigen is usually the costliest factor of an immunoassay. The price of Spike RBD in the market can range between US$ 2,000-5,000 per mg https://www.abbexa.com/sars-cov-2-spike-protein-rbd-l452r-mutation (accessed 10/12/2021).

The high cost of production relies on the fact that Spike RBD is obtained using eukaryotic protein expression platforms. Even though cheaper eukaryotic expression systems have been developed including the use of yeast *Pichia pastoris* [9]. The price of production is still high in comparison to proteins produced in prokaryote expression platforms such as *Escherichia coli* [10].

The fact that the SARS-CoV-2 Spike protein is highly glycosylated and contains several disulfide bridges makes this antigen difficult to obtain in *E coli*. The lack of glycosylation systems and the reducing environment typically found in the bacterium cytoplasm results in formation of insoluble inclusion bodies upon expression of Spike in *E. coli* [11]. Some researchers have shown that Spike RBD can be obtained in soluble form from *E. coli* after a denaturation/refolding procedure and/or by using genetically modified strains with reducing cytoplasm [12], [13], [14], [15], [16]. In any case, in all studies were Spike RBD has been obtained from *E. coli* the antigen seems to have limited antigenicity and low yield [12]. This may explain why there is no report of an accurate immunoassay, validated with a large cohort of samples, which uses Spike RBD produced in *E. coli*.

We have described previously an ultrafast, simple, and inexpensive Ni^2+^ magnetic bead immunoassay which allows detection of human IgG reactive to SARS-CoV-2 nucleocapsid protein using a minimal amount of sample and delivering the results in less than 7 min [17], [18]. The principle of the assay relies on quick and direct mobilization of 6xHis tagged version antigens to the surface of Ni^2+^ magnetic beads. We demonstrated that this assay can be used to detect human antibodies reactive to SARS-CoV-2 full length prefusion Spike and to Spike RBD which were recombinantly produced in eukaryotic expression systems.

Here we demonstrate that our previously described assay can also be used to identify human IgG reactive to a fragment of SARS-CoV-2 Spike containing the RBD which was recombinantly produced in *E. coli*.

## Materials and Methods

### Expression and purification of recombinant receptor-binding domain of SARS-CoV-2 spike protein

A codon optimized synthetic gene was produced and cloned into the pET28a vector by General Biosystems. The plasmid construction was named pLHSarsCoV2-S and allowed the expression of a N-terminal 6xHis tagged fusion of a fragment of SARS-CoV-2 Spike S1 containing part of the N-terminal domain and the full RBD (Uniprot MN908947, M153 to T589). The plasmid construction was transformed into *E. coli* BL21 (λDE3) and the cells were grown in 100 ml LB medium containing kanamycin (50 µg.ml^-1^) at 120 rpm at 37°C to an OD_600nm_ of 0.3. The incubator temperature was changed to 16°C, after 30 min, IPTG was added to a final concentration 0.3 mM. The culture was kept at 120 rpm at 16° over/night. Cells were collected by centrifugation at 3,800 xg for 5 min. The cell pellet was resuspended in 10 ml of buffer 1 (Tris-HCl pH 8 50 mM, KCl 100 mM, Urea 8 M). Cells were disrupted by sonication on an ice bath with 8 min cycle, 15 sec pulse and 15 sec rest and amplitude 35%. The soluble fraction was recovered after centrifugation at 11.000 rpm for 10 minutes at 4°C and loaded into a 5 ml Histrap chelating Ni^2+^ column (Cytiva) which has been previously equilibrated with buffer 1. The column was washed with 20 ml of buffer 1 and bound proteins were eluted with 5 ml of buffer 1 with increasing imidazole 50 mM, 100 mM, 300 mM and 500 mM. Fractions of 1 ml were collected and analyzed by SDS-PAGE. The fractions containing the protein of interested were polled and the protein was stored in aliquots at - 20°C.

### Human samples

Human samples were collected at Hospital Erasto Gaertner in Curitiba and Federal University of Paraná in Matinhos. Samples for serological analysis comprised both serum and plasma-EDTA. COVID-19 positive cases were confirmed by the detection of SARS-CoV-2 RNA via real-time RT-PCR from nasopharyngeal sample swabs. The time point of sampling of serum ranged from 1 to 100 days after PCR detection. Among the 86 COVID-positive cases there were 7 convalescents being 1 asymptomatic and 6 mild non-hospitalized cases. All remaining samples were collected within the first 14 days of the hospitalization period and included 39 severe and 40 critical (intensive care unit). The cohort of 204 negative controls consisted of pre pandemic samples collected in 2018. The Institutional Ethics Review Board CEP/HEG (n# 31592620.4.1001.0098 and n# 54095221.0.0000.0098), CEP/UFPR (n# 43948621.7.0000.0102 and n# 35872520.8.0000.0102) approved this study. Informed consent was obtained from all participants in this study. All methods were performed in accordance with the relevant guidelines and regulations.

### Magnetic beads-based immunoassay

The magnetic bead-based immunoassay was developed using Ni^2+^ magnetic beads as described previously with few modifications [18]. One milliliter of Ni^2+^ magnetic particles MagneHis (Promega cat V8550) was transferred to a 50 ml Falcon tube and washed twice with 1 ml of TBST 1X. The resin was resuspended in 25 ml of TBST 1X and 4 mg of purified N-terminal 6x His tagged Spike-RBD protein was added. After mixing by inversion, TBST 1X was added to a final volume of 50 ml. The mixture was incubated for 10 min at room temperature with mixing by inversion every 2 min. Beads were washed with 25 ml of TBST 1X and finally resuspended in 5 ml of TBST 1X and stored at 4° C for up to 2 months.

Assays were performed in 96-well flat-bottom polystyrene microplates (OLEN). An aliquot of 0.8 ml of antigen loaded beads were resuspended in 10 ml of TBST 1X containing skimmed milk 1% (w/ v). After thoroughly mixing, 0.1 ml of the beads were added to each well of the first 96 well plate. The second plate was prepared by adding 0.2 ml of TBST 1X containing skimmed milk 1% (w/ v) and 4 µl of samples or controls to each well. The wells of plates 3 and 4 contained 0.2 ml of TBST 1X and urea 1 mol/l. Plate 5 contained 0.15 ml of goat anti-human IgG-HPR (Thermo Scientific) at 1/1,500 dilution in TBST 1X. The wells of plates 6 and 7 received 0.2 ml of TBST 1X and plate 8 received 0.15 ml of HPR chromogenic substrate TMB (Thermo Scientific).

When all plates were set, reactions started by transferring the beads from plate 1 to 8 and the applying the following incubation times: plates 2 and 5 (2 min); plates 3, 4, 6 and 7 (30 sec); plate 8 (8 min). Beads were gently mixed during incubation. Beads transfer and homogenization was achieved using home-made manually operating magnetic transfer and homogenization device described previously. After completion of the reaction on plate 8, beads were removed, the OD_650nm_ was measured using a Tecan M Nano microplate reader (TECAN) with a bandwidth of 9 nm and 25 flashes.

## Data analysis

One COVID-19 positive serum was used as reference throughout the study. Raw data were normalized as % of this reference before applying Receiver Operating Analysis (ROC) using GraphPad Prism 7.0. Statistical analysis was performed using the t test on GraphPad Prism 7.0.

## Results and Discussion

We have described previously a nickel magnetic bead immunoassay which was successfully applied to track SARS-CoV-2 seroconversion in humans using either prokaryotic produce nucleocapsid or eukaryotic produced full-length Spike or Spike RBD as antigens [18]. The assay is based on the use of commercially available Ni^2+^ magnetic particles which can be directly coated with purified 6x His-tagged SARS-CoV-2 antigens. The method is amenable to high throughput and can process samples 96 samples delivering ultrafast results. Despite the simplicity and low cost of the assay one limiting factor is the price of the Spike and Spike RBD antigens which can be found from 2,000 to 5,000 US$/mg on the market.

Previous studies indicated that fragments of the SARS-CoV-2 Spike can be expressed at high levels using *E. coli* as host [13]. However, these constructions are typically produced as insoluble inclusion bodies. Even though soluble protein can be obtained after denaturation/refolding procedures the antigenicity of these constructions seems to be limited [13], [15]. To the best of our knowledge, there is no commercially available immunoassay based on the use of Spike antigen produced in *E. coli*.

We obtained a synthetic gene which was able to express a fragment of the Spike S1 protein containing part of the N-terminal domain and the full length RBD from the T7 promoter using *E. coli* as expression vector. As expected, based on previous studies, this Spike construction was found as insoluble inclusion bodies after expression [13]. All attempts to reduce protein aggregation which included protein induction at 16°C, use of detergents during cell disruption and co-expression with the cysteine reducing system CysDiCo; were unsuccessful in solubilizing the Spike antigen (data not shown) [12].

To purify the Spike construction, the *E. coli* cell pellet obtained after protein induction was directly resuspended in buffer containing urea 8 mol/l. Cells were disrupted by sonication and the 6x His tagged Spike S1 RBD was purified using Ni^2+^ affinity chromatography using imidazole as eluent, all steps were performed in the presence of urea 8 mol/l. The Spike S1 RBD could be purified 90% homogeneity (Fig. 1) and the protein yield was 120 mg/l of cell culture which is way higher than typically obtained in eukaryotic systems. The estimated cost of consumables for antigen production was US$ 9 US$/mg which is 15 x below the price for production of pre fusion Spike in HEK cells [7].

**Fig. 1.**
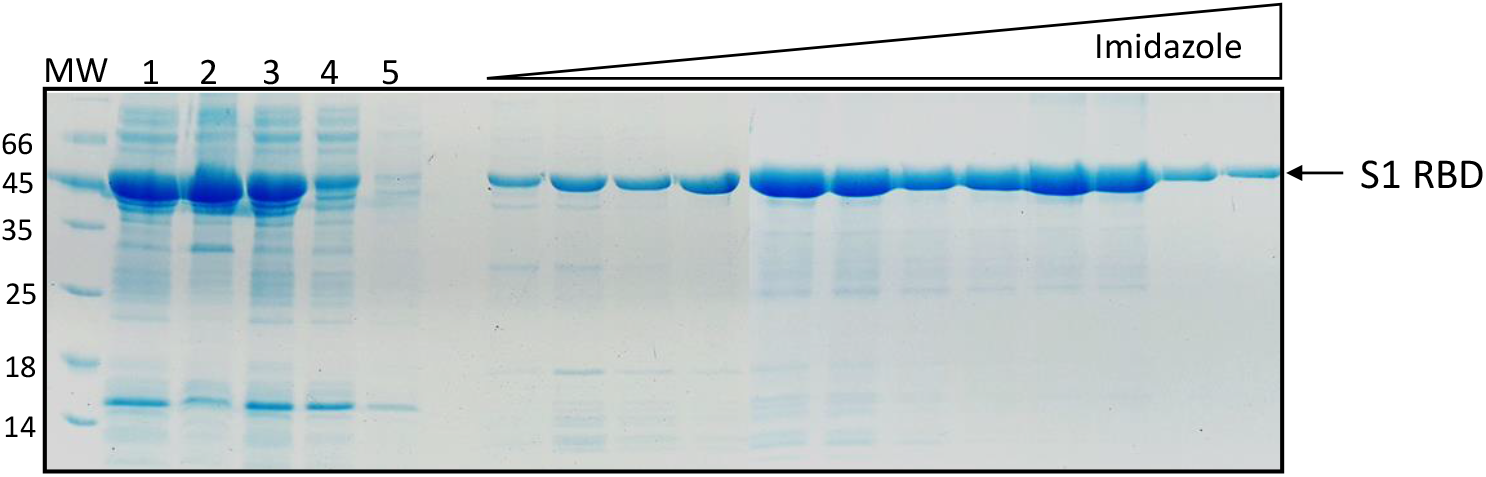
Purification of *E. coli* induced Spike S1 RBD. SDS-PAGE analysis of the fractions eluted from the Hitrap chelating column. Lane 1, whole cell extract; lane 2, insoluble fraction; lane 3, soluble fraction; lane 4, flow through; lane 5, wash buffer 1. Other lanes, fractions collected of the imidazole gradient from 50, 100, 300 to 500 mM. The Spike S1 RBD is indicated by an arrow and eluted between 100- and 500-mM imidazole.

The Spike S1 RBD antigen obtained after affinity chromatography was mobilized to the surface of Ni^2+^ magnetic beads. The Spike protein preparation containing urea was incubated with the nickel beads and diluted in TBST buffer in such way that the bead coating step also acted as Spike refolding step. The development of the immunoassay was based on our previous described method [12]. The investigated serum was incubated with the beads for 2 min; followed by 2x 30 sec washing steps; 2 min incubation with anti-human IgG HPR conjugate; 2x 30 sec washing and final 8 min incubation in chromogenic HPR substrate (TMB) before the measurement of OD_650nm_ using a microplate reader. The complete procedure takes about 15 min.

A cohort of 28 pre-pandemic and 14 qRT-PCR COVID-19 confirmed serum samples were initially evaluated to determine the best assay conditions. This initial screening revealed that the negative cohort exhibited high background (mean OD_650nm_ = 0.13) and they were not clearly distinguished from the COVID-19 positive serum samples (Fig. 2A). Reports on the literature indicate that washing buffers containing urea can be used to reduce the background of negative serum in classic ELISA assays [19], [20]. In fact, use of urea in ELISA washing buffers is a well-established method to detect high affinity antibodies. Only antibodies exhibiting high affinity will remain bound to the antigen in the presence of the chaotropic agent. Previous studies indicate that the addition of urea into ELISA wash buffers can increase the specificity of the assay enabling the distinction between closely related diseases such as Zika and Dengue [19], [20].

**Fig 2.**
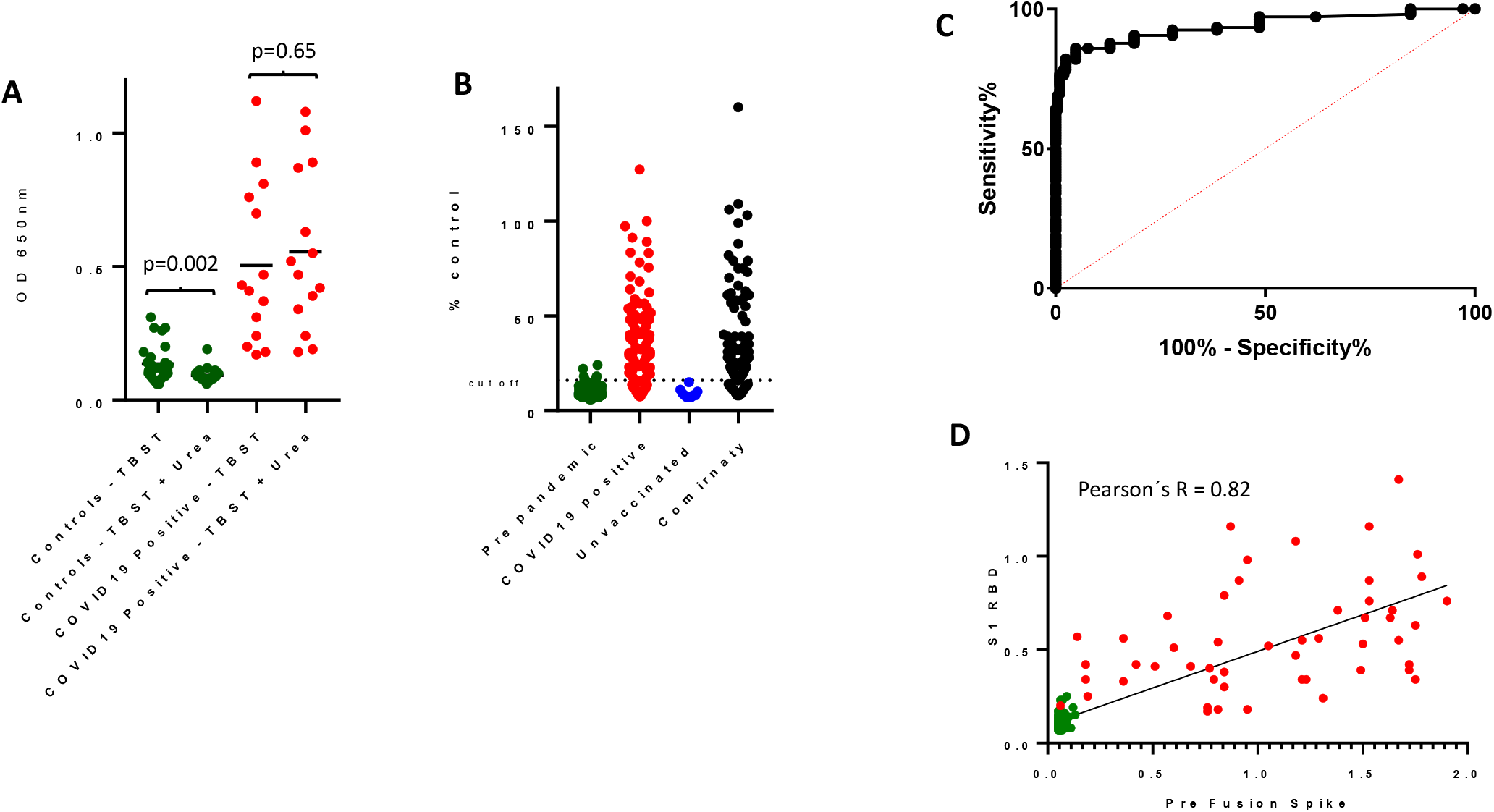
Development of the assay to detect human IgG reactive to S1 RBD expressed in *E. coli*. A) Comparison of raw OD_650nm_ obtained with regular TBST buffer or TBST containing urea 1 mol/l. Pre-pandemic samples are shown in green, qRT-PCR COVID-19 confirmed cases in red. B) Validation of the assay was performed analyzing IgG levels to S1 RBD in pre-pandemic (green) and qRT-PCR COVID-19 confirmed cases sera. The assay was also able to detect IgG in sera obtained from Comirnaty vaccinated individuals (black), unvaccinated (blue). The assay cut off at >98% specificity is indicated by the dashed line. C) ROC curve analysis of the developed assay. D) The levels of IgG (raw signal OD_600_) reactive to S1 RBD obtained from *E. coli* was well correlated with the levels detected using Spike obtained from HEK293-3F6 cells as antigen (Pearson’s R = 0.82, p <0.0001).

We evaluated if addition of urea to the wash buffers could reduce the background signal of the negative cohort. We noted that the addition of urea 1mol/l to the wash buffer significantly reduced the background of the negative serum cohort (mean OD_650nm_ = 0.09, p=0.0024) without reducing the signal obtained from the positive serum cohort (Fig. 2A). Hence, urea 1mol/l was added to the buffers in the two consecutive 30 sec wash steps after beads incubation with the investigated serum.

After setting the assay conditions the method was further validated using a larger cohort. The presence of IgG reactive to SARS-CoV-2 Spike S1 RBD antigen was analyzed in sera from 208 pre pandemic samples and 86 qRT-PCR COVID-19 confirmed cases. The pre-pandemic control sera produced only background signal which were clearly distinguished from the RT-qPCR-confirmed COVID-19 cases (Fig. 2B). Receiver operating characteristic (ROC) analysis revealed an area under curve (AUC) of 0.94 (Fig. 2C). A sensitivity of 82% (73 to 89 % at 95% CI) could be achieved at a cost of 98% (94 to 99 % at 95% CI) specificity. These number indicate that the method operate with good accuracy. The biological relevance of the developed assay was further confirmed by the fact the levels of IgG reactive to SARS-CoV-2 Spike S1 RBD antigen produced in *E. coli* was well correlated with the levels of IgG reactive to prefusion Spike produced in HEK cell lines (Pearson’s R = 0.82, Fig. 2D).

We determined if the developed method could be used to detect human antibodies raised after vaccination. Of the fifteen samples collected from unvaccinated subjects which reported to have not developed COVID-19, all samples remained below the assay cut off value (Fig. 2B). On the other hand, of the 120 samples collected from subjects fully immunized with Comirnaty (and reported to have not developed COVID-19), 85% were positive (Fig. 2B). These data indicate that our assay can not only detect high affinity IgG reactive towards Spike S1 RBD which were raised from previous SARS-CoV-2 infections but also those developed after vaccination with good precision and at low cost.

In conclusion, here we show that *E. coli* expression system can be used to produce a N-terminal 6x His tagged SARS-CoV-2 Spike S1 RBD that can be purified at low cost and high yield. The protein retained its antigenic properties when refolded on the surface of nickel magnetic particles and could be used detect high affinity IgG present in sera from COVID-19 positive cases and from individuals fully immunized with Comirnaty. The method operated with good accuracy, high throughput, and low cost (less than US$ 1/per sample). To the best of our knowledge this is the first report to show the feasibility of using Spike produced in *E. coli* to develop an accurate immunoassay which has been subject to validation with more than 100 samples. We believe that this novel method may act as an important toll to identify and quantify high affinity IgG in human samples reactive to Spike RBD in large scale populational studies.

## Data Availability

All data produced in the present study are available upon reasonable request to the authors

## Author’s Contributions

L.F.H conceived and designed the study. L.F.H, M.S.C, A.C.G, N. M. P, F.G.M.R, M.N.A, D.L.Z and J.M.N collected the samples. L.F.H, M.S.C and A.C.G analyzed the data and wrote the paper.

## Notes

Federal University of Paraná UFPR has filled patent protections for nickel magnetic immunoassay process, magnetic COVID-19 immunological test product and magnetic bead extractor device. All designed product and processes will be freely available for academic and non-commercial users.

## Acknowledgment

We are grateful to Prof. Karl Forchhammer (Tubingen University / Germany) for initial support for this project. This work was supported by the Alexander von Humboldt foundation, UFPR, CNPq, CAPES and by the Fundação Araucária.

## References

1. Chavez S, Long B, Koyfman A, Liang SY. Coronavirus Disease (COVID-19): A primer for emergency physicians. Am J Emerg Med. 2021;44:220–229. doi:10.1016/J.AJEM.2020.03.036

2. Lan J, Ge J, Yu J, et al. Structure of the SARS-CoV-2 spike receptor-binding domain bound to the ACE2 receptor. Nat 2020 5817807. 2020;581(7807):215–220. doi:10.1038/s41586-020-2180-5

3. Weisblum Y, Schmidt F, Zhang F, et al. Escape from neutralizing antibodies by SARS-CoV-2 spike protein variants. Elife. 2020;9:1. doi:10.7554/ELIFE.61312

4. Dispinseri S, Secchi M, Pirillo MF, et al. Neutralizing antibody responses to SARS-CoV-2 in symptomatic COVID-19 is persistent and critical for survival. Nat Commun. 2021;12(1). doi:10.1038/S41467-021-22958-8

5. Márquez-Ipiña AR, González-González E, Rodríguez-Sánchez IP, et al. Serological Test to Determine Exposure to SARS-CoV-2: ELISA Based on the Receptor-Binding Domain of the Spike Protein (S-RBDN318-V510) Expressed in Escherichia coli. Diagnostics 2021, Vol 11, Page 271. 2021;11(2):271. doi:10.3390/DIAGNOSTICS11020271

6. Krähling V, Halwe S, Rohde C, et al. Development and characterization of an indirect ELISA to detect SARS-CoV-2 spike protein-specific antibodies. J Immunol Methods. 2021;490:112958. doi:10.1016/J.JIM.2021.112958

7. Alvim RGF, Lima TM, Rodrigues DAS, et al. An affordable anti-SARS-COV-2 spike protein ELISA test for early detection of IgG seroconversion suited for large-scale surveillance studies in low-income countries. medRxiv. September 2020:2020.07.13.20152884. doi:10.1101/2020.07.13.20152884

8. Perkmann T, Perkmann-Nagele N, Koller T, et al. Anti-Spike Protein Assays to Determine SARS-CoV-2 Antibody Levels: a Head-to-Head Comparison of Five Quantitative Assays. Microbiol Spectr. 2021;9(1). doi:10.1128/SPECTRUM.00247-21/ASSET/D37158B1-1196-41B7-9587-185BC91C7821/ASSETS/IMAGES/LARGE/SPECTRUM.00247-21-F004.JPG

9. Consortium AA. Structural and functional comparison of SARS-CoV-2-spike receptor binding domain produced in Pichia pastoris and mammalian cells. Sci Reports 2020 101. 2020;10(1):1–18. doi:10.1038/s41598-020-78711-6

10. Villafañe L, Vaulet LG, Viere FM, et al. Development and evaluation of a low cost IgG ELISA test based in RBD protein for COVID-19. J Immunol Methods. 2021;500:113182. doi:10.1016/J.JIM.2021.113182

11. Prahlad J, Struble LR, Lutz WE, et al. Bacterial expression and purification of functional recombinant SARS-CoV-2 spike receptor binding domain. bioRxiv. February 2021:2021.02.03.429601. doi:10.1101/2021.02.03.429601

12. Prahlad J, Struble LR, Lutz WE, et al. CyDisCo production of functional recombinant SARS-CoV-2 spike receptor binding domain. Protein Sci. 2021;30(9):1983–1990. doi:10.1002/PRO.4152

13. Fitzgerald GA, Komarov A, Kaznadzey A, Mazo I, Kireeva ML. Expression of SARS-CoV-2 surface glycoprotein fragment 319–640 in E. coli, and its refolding and purification. Protein Expr Purif. 2021;183:105861. doi:10.1016/J.PEP.2021.105861

14. He Y, Qi J, Xiao L, Shen L, Yu W, Hu T. Purification and characterization of the receptor_binding domain of SARS_CoV_2 spike protein from Escherichia coli. Eng Life Sci. 2021;21(6):453. doi:10.1002/ELSC.202000106

15. Bellone ML, Puglisi A, Dal Piaz F, Hochkoeppler A. Production in Escherichia coli of recombinant COVID-19 spike protein fragments fused to CRM197. Biochem Biophys Res Commun. 2021;558:79. doi:10.1016/J.BBRC.2021.04.056

16. Maffei M, Montemiglio LC, Vitagliano G, et al. The nuts and bolts of SARS-CoV-2 spike receptor-binding domain heterologous expression. Biomolecules. 2021;11(12). doi:10.3390/biom11121812

17. Huergo LF, Selim KA, Conzentino MS, et al. Magnetic Bead-Based Immunoassay Allows Rapid, Inexpensive, and Quantitative Detection of Human SARS-CoV-2 Antibodies. ACS Sensors. 2021;6(3):703–708. doi:10.1021/ACSSENSORS.0C02544/SUPPL_FILE/SE0C02544_SI_001.PDF

18. Conzentino MS, Santos TPC, Selim KA, et al. Ultra-fast, high throughput and inexpensive detection of SARS-CoV-2 seroconversion using Ni2+ magnetic beads. Anal Biochem. 2021;631:114360. doi:10.1016/J.AB.2021.114360

19. Tsai WY, Youn HH, Tyson J, et al. Use of Urea Wash ELISA to Distinguish Zika and Dengue Virus Infections. Emerg Infect Dis. 2018;24(7):1355. doi:10.3201/EID2407.171170

20. Wang Q, Lei Y, Lu X, et al. Urea-mediated dissociation alleviate the false-positive Treponema pallidum-specific antibodies detected by ELISA. PLoS One. 2019;14(3). doi:10.1371/JOURNAL.PONE.0212893

